# Real-time outlier detection in digital PCR data for wastewater-based pathogen surveillance

**DOI:** 10.1101/2025.09.30.25336068

**Authors:** Rachel E. McLeod, Timothy R. Julian, Tanja Stadler, Adrian Lison

## Abstract

Wastewater-based epidemiology provides insights into the spread of infectious diseases by sampling and analyzing wastewater from wastewater treatment plant catchments. Longitudinal measurements of pathogen concentrations in wastewater have been shown to align with disease incidence and can be used to estimate indicators of epidemic growth such as effective reproduction numbers (R_*t*_). This informs public health decision-making by providing feedback on the transmission of disease and effectiveness of interventions. However, methods for analysing growth trends from wastewater are sensitive to multiple sources of variation during shedding, in-sewer transport, sample collection, and processing. A particular challenge is posed by occasional but high outlier measurements, which can bias estimated trends and epidemiological parameters during seasonal and epidemic waves. The identification and handling of such data points in real-time wastewater monitoring is important to provide a reliable basis for public health decision-making. In this study we propose a fast and simple outlier detection method for digital PCR measurements from wastewater that allows identification of outliers in real time. We evaluated our method by assessing its potential to improve real-time trend estimates. We show that, in the majority of situations, the removal of an outlier identified by our method makes the real-time trend estimate for a particular time point more representative of the trend which is retrospectively obtained for the same time point. Our method provides a simple way for laboratories to flag potential outliers in time series data, improving data quality for further analysis.

## 1. Introduction

Wastewater-based epidemiology (WBE) uses samples collected from municipal wastewater to infer relative prevalence and transmission of an infectious disease within the population of a wastewater catchment [1]. This is achieved by measuring the concentration and total load of bacterial or viral gene copies in the wastewater using molecular techniques such as polymerase chain reaction (PCR) [2, 3, 4]. Pathogen loads in wastewater can give an indication of the prevalence of the pathogen in the population and have been shown to correlate with clinical case data for COVID-19 and other diseases [5, 6, 7]. Moreover, trends in wastewater load over time can be used to predict hospitalisations and ICU admissions [8, 9] and to estimate the effective reproduction number R_*t*_ [6, 10, 11, 12, 13, 14].

However, wastewater data are subject to multiple uncertainties and sources of variation, including fluctuating population sizes of wastewater treatment plant catchments, unknown and uneven shedding of infected individuals, variable sampling from sewage, degradation of genetic material in the sewer and during sample transport, and varying extraction efficiency and PCR inhibition in the lab [15, 16]. These factors make it difficult to relate wastewater measurements to absolute numbers of infected individuals in the sampled catchment, and to reliably track trends in infection incidence. Thus, analyses of WBE data typically apply smoothing to measured pathogen concentrations or loads [17]. Similarly, many public wastewater dashboards show smoothed load trends, for example using a centrally-aligned rolling average [18, 19, 20].

Although smoothing methods can effectively reduce noise in WBE data, they often remain susceptible to single extreme measurements that strongly deviate from the overall trend for no apparent reason. Such measurements are regularly observed in WBE data from different countries, affecting statistical analyses and real-time monitoring, and are often referred to as outliers [21, 2, 22, 15]. Some outliers may be eliminated through appropriate laboratory quality control, for example using controls to flag samples with low extraction efficiency or increased PCR inhibition [23, 2]. When data are subject to such quality control our confidence in their accuracy as a measure of the RNA concentration in the sample increases, leading us to assume that the observed outliers in the data set accurately represent the wastewater sample’s viral concentration. Note that this does not directly translate to an assumption of an accurate measure of viral load in the wastewater catchment, rather we attribute the extreme value to other sources of variation which cannot be identified or controlled for with current laboratory quality control. The need to identify extreme measurements whose estimated viral loads may not accurately reflect disease dynamics makes outlier detection a crucial step in the analysis of WBE data [24].

To detect outliers from wastewater time series, researchers have employed various methods, such as identifying days with anomalously high or low flow [25, 26], flagging large deviations from a fitted trend line [27, 28], or fitting autoregressive models [29]. However, these methods make simplistic assumptions about the variability of wastewater measurements or rely on historical data to define an expected or typical level of variability. Some wastewater dashboards have also adopted more robust smoothing approaches, for example using a rolling median or trimmed mean where maximum and minimum values in the rolling window are removed. However, these approaches are mostly suitable for retrospective smoothing of wastewater concentrations, and can bias trend estimates when applied to the most recent observations in a real-time fashion.

In this paper, we propose a method that detects outliers in real time with little historical data by using knowledge about the measurement noise in wastewater data. The method is tailored to pathogen concentrations measured via digital PCR (dPCR), a technique that quantifies nucleic acids by partitioning the sample into many individual reactions and counting the number of positive partitions. Our approach takes as input key laboratory parameters of a dPCR assay, including the dilution factor and the number and volume of partitions, which are readily available to the reporting laboratory or can be derived from standard lab protocols. Using this information, we apply an accurate model of dPCR-related measurement noise to identify abnormal levels of variability in wastewater measurements without the need for extensive calibration. We have tailored our method specifically to detect “high outliers”, *i.e*. measurements that lie considerably above the expected trend. In contrast to “low outliers”, *i.e*. measurements below the expected trend, high outliers are not bounded and can reach extreme values, as has been observed in WBE data from multiple countries [21, 2, 22, 15]. When unaccounted for, high outliers can strongly bias trend estimates and affect the real-time interpretation of wastewater measurements, especially during seasonal or epidemic waves of the monitored pathogen.

To evaluate the potential of our method for improving the robustness of real-time trend estimates by detecting and removing high outliers, we apply it to three years of wastewater measurements for four different respiratory viruses at fourteen locations in Switzerland. We show that, even for pathogens with limited historical data, our method reliably detects outliers that, if left uncorrected, would lead to substantial deviations in trend estimation. This enables laboratories to flag and report outliers as soon as new measurements become available, improving real-time analyses and visualisation on monitoring dashboards.

## 2. Methodology

### 2.1. Outlier detection method

The method to identify outliers consists of six steps (overview in Figure 1), which we describe along with definitions of parameters and explanations of underlying assumptions.

**Figure 1:**
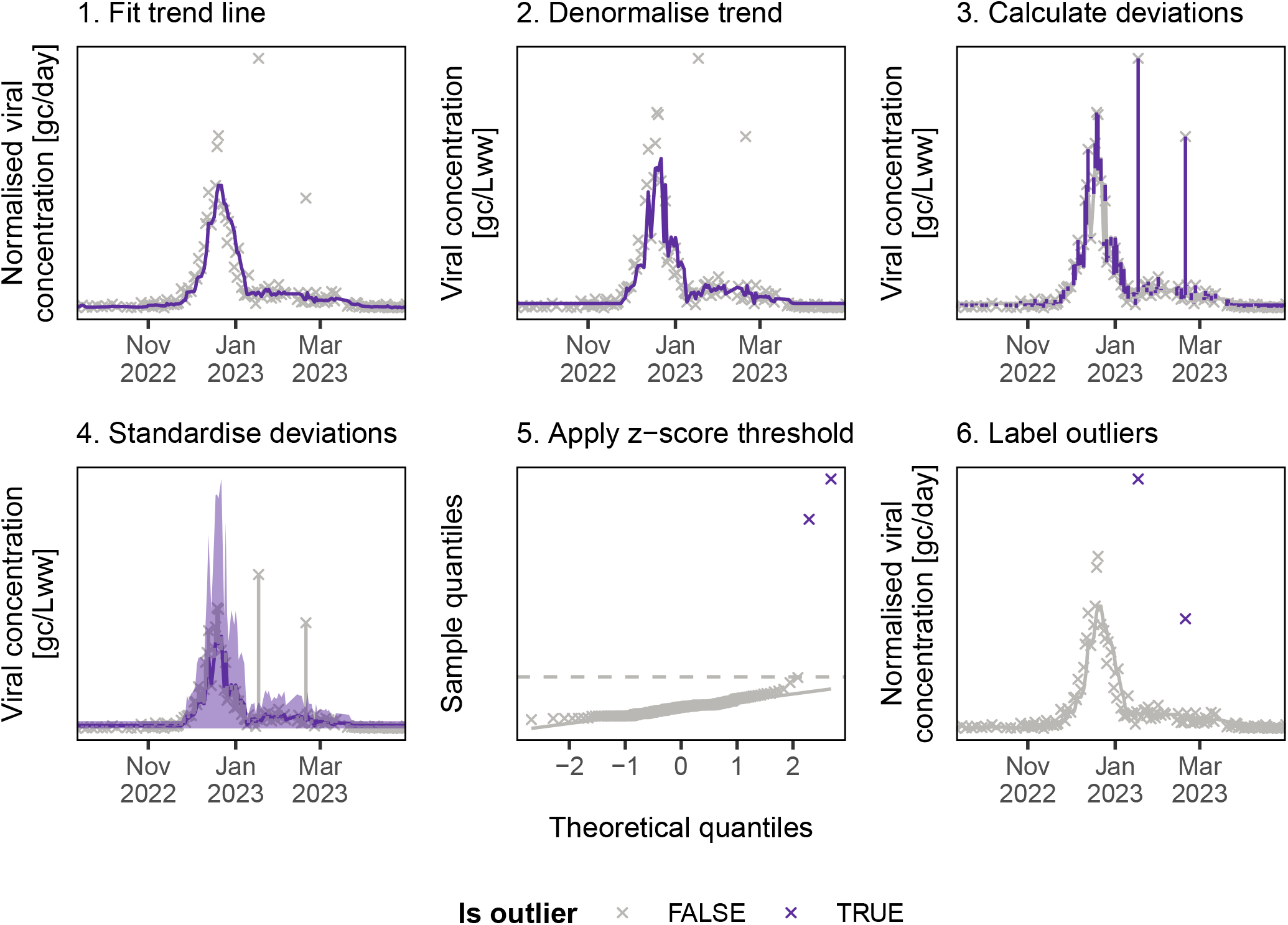
Overview of the outlier detection method. The method consists of six steps which are repeatedly applied to identify and filter outliers from the data set until no further outliers are found. 1) A trend line is fitted through normalised viral concentration measurements. 2) This trend is denormalised back to a concentration trend. 3) All deviations between concentration measurements and the concentration trend are calculated. 4) The deviations are standardised by dividing by the expected standard deviation at the concentration trend. 5) A z-score threshold of three is applied. 6) Any data points above the threshold are labeled as an outlier.

The digital PCR (dPCR) assay measures the viral RNA concentration, *c*_*t*_, in the wastewater for a particular day, *t*. To reduce variation in the time series from differences in flow through the sewage system, we calculate a normalised concentration, ĉ_*t*_, as

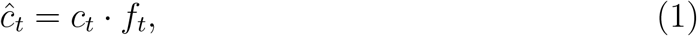

where *f*_*t*_ is the flow through the wastewater treatment plant (WWTP) for day *t*. Although we here focus on flow-based normalization, other normalisation approaches, e.g. using faecal markers such as CrAssphage or PMMoV [30], could be applied in an analogous fashion.

We then fit a trend line, T(ĉ_*t*_), on the normalised concentrations ĉ_*t*_ using a right-aligned weighted rolling median over a window of *k* days, *i.e*.

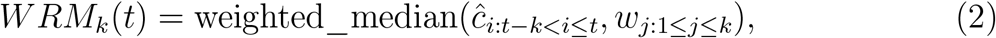

with *k* = 7, weights *w* subject to 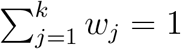 and the weighted_median defined as the normalised concentration at time *i* that satisfies

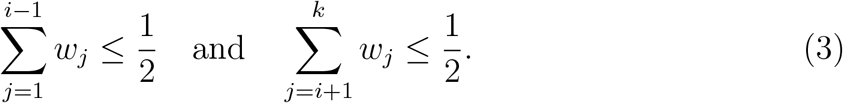

Linearly decreasing weights 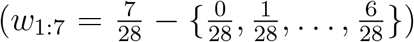 are given to observations, with most recent observations receiving the highest weight. Missing data points and their corresponding weights are removed before calculating T(ĉ_*t*_) = *WRM*_*k*_(*t*).

To identify outliers, we assume that E[ĉ_*t*_] ≈ T(ĉ_*t*_) for non-outlier measurements, such that large deviations from this expectation suggest the presence of an outlier. However, this requires adjusting for the expected, i.e. “normal”, measurement variation given the sample concentration on day *t*. For this, we de-normalise the concentration trend T(ĉ_*t*_) as

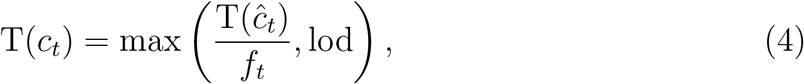

*i.e*. we divide the normalised concentration trend T(ĉ_*t*_) by the daily wastewater flow, avoiding zero values by truncation with the limit of detection (lod) of the dPCR assay. We then predict the coefficient of variation for non-outlier dPCR measurements on each date *t*, defined as [31]

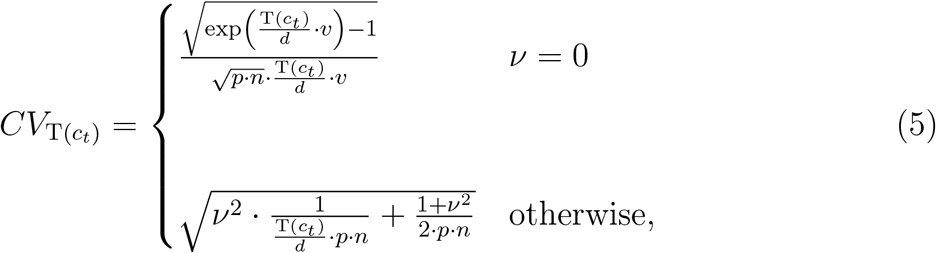

where *n* is the number of technical replicates per sample, *p* is the (average) number of partitions in the dPCR reaction, *v* is the partition volume, *d* is the dilution factor of the sample, and ν is the pre-PCR coefficient of variation (see Section 2.2.2 for explanations of these variables).

To obtain a measure of the spread of observations around the trend, we calculate the standardised deviation of the measured concentrations from T(*c*_*t*_),

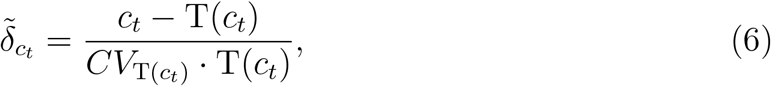

where 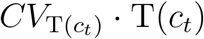 is the standard deviation of non-outlier measurements as predicted by equation 5. We consider the resulting standardised deviations 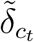 as standard normal distributed, *i.e*. 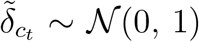, which holds approximately true unless concentrations are close to zero or approach the saturation of the dPCR assay. Under this assumption, we can label a measurement *c*_*t*_ as an outlier by applying a z-score threshold of three [32, 33], *i.e*.

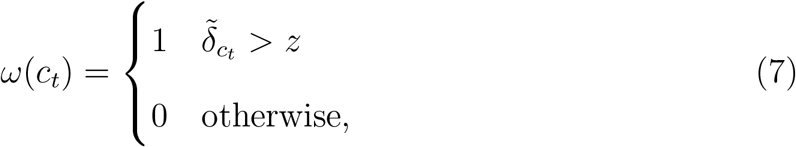

where *ω*(*c*_*t*_) = 1 represents a high outlier observation (deviation above the trend), *ω*(*c*_*t*_) = 0 a non-outlier observation, and *z* = 3 corresponds to the 99.9% quantile of the standard normal distribution. We iteratively apply this method by filtering identified outliers from the data set and repeating the above steps until no further outliers are found.

### 2.2. Evaluation of method performance

Here we detail the data set used and approach taken for evaluating the performance of our method. We conducted a small analysis for situations where per-sample metadata are unavailable and describe the implementation in R.

#### 2.2.1. Wastewater time series data

To evaluate our method on real-world data, we used publicly available wastewater measurements provided by the Wastewater Monitoring Laboratory at Eawag, the Swiss Federal Institute of Aquatic Science and Technology [34]. These data were obtained using dPCR to quantify viral RNA extracted from wastewater samples collected at various locations throughout Switzerland (Table S1). On average, four 24-hour composite wastewater samples per week and WWTP were processed for viral RNA extraction, concentration and quantification.

In total, the wastewater data contained several years of measurements for five viral targets (SARS-N1, SARS-N2, IAV-M, IBV-M, RSV-M/N), from fourteen WWTPs (Table S1), resulting in 70 time series of varying lengths which were available for method testing and evaluation (Figure S1). Since December 2021 SARS-CoV-2 viral concentrations have been measured for six WWTPs using an assay targeting the N1 protein gene of the SARS-CoV-2 virus and Murine Hepatitis Virus as internal control (*N1MHV* assay) [6]. In November 2022 an additional four targets were added to the monitoring programme using an assay for Influenza A and B, Respiratory Syncytial Virus, and SARS-CoV-2 targeting the N2 protein gene (*RESPV4* assay) [11]. In July 2023, these two assays were combined into a single *RESPV6* assay [2] and an additional eight WWTPs were added to the monitoring programme. In January 2025 monitoring was reduced from fourteen to ten WWTPs. All reported measurements passed quality control [2] and were normalised to account for daily flow volumes in litres (L) at the WWTP by converting concentration measurements of viral gene copies per litre of wastewater (gc/Lww) into normalised concentrations of viral gene copies per day (gc/day).

#### 2.2.2. Laboratory parameters

In addition to viral concentration and flow measurement data, we obtained details of the laboratory protocols, which were used for parameterising the dPCR model in equation 5. Mainly these details relate to conditions from the dPCR experiment including: a dilution factor, which represents how much the original sample was diluted during processing and is a combination of various volumes used in the extraction and dPCR protocol (see Appendix A), generally 30 or 50 for our data set; the number of replicates, which is the number of technical replicates of a single sample run on the dPCR assay, generally two; the total partitions of the dPCR reaction, on average around 22,000 [15,000 - 30,000]; and the partition volume, which is the volume of a single dPCR partition and is dependent on the dPCR reaction reagents and system, 0.519 nL. Additionally, the pre-PCR coefficient of variation, ν, represents sources of variation before the PCR reaction, including variation in sewage characteristics, sampling collection and laboratory extraction and preprocessing, which are often difficult to quantify. For our application, however, it is sufficient to choose ν such that the standardised trend deviations 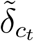 have a variance of approximately one. For this, we computed the median standard deviation of the 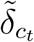over all treatment plants for each viral target at different values of ν ranging from 0 to 1 in steps of 0.01, and selected the value of ν for which the median standard deviation was closest to one. This yielded pre-PCR coefficients of variation ranging between 0.46-0.64 across the different targets (Figure S2).

#### 2.2.3. Evaluation approach

To assess whether our outlier detection method can improve real-time trend estimates by excluding high outliers, we evaluated the difference between real-time trend estimates with and without outliers in comparison to a robust retrospective trend estimate (*i.e*. when all data were available), which is generally less sensitive to occasional single outliers than real-time estimates. An advantage of this evaluation approach is that it does not require a “ground truth” labeling of outliers and allows us to consider the varying magnitudes of distortion that outliers can have on the estimated trend.

Real-time trends were estimated on each date with new observations, using loess with least-squares fitting [35], which is a common smoothing method used in modeling of WBE data [12, 13]. A retrospective trend was also obtained using loess, however, here we used four iterations of an M-estimator using Tukey’s biweight function, which provides more robust smoothing. When estimating the real-time trend with outlier removal, we replaced outliers detected by our method by linear extrapolation of the trend at the most recent non-outlier observation.

We compared the real-time trend estimates with and without outlier removal against the retrospective trend by computing the mean absolute scaled error

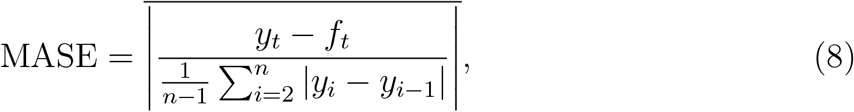

where *y*_*t*_ is the retrospective trend at time *t, f*_*t*_ is the real-time trend either with or without outliers removed and *n* is the number of observations in the time series. We used the sign test to test for consistent differences in MASE scores between real-time trends with and without outlier removal.

#### 2.2.4. Fixed parameters analysis

To account for settings where detailed per-sample metadata are not available for the laboratory parameters, we repeated our outlier analysis fixing these parameters to set values. The number of replicates and dilution factor were fixed at the most common values of 2 and 30 respectively. The number of partitions was fixed at the instrument-specific maximum of 30,000 partitions. The pre-PCR variation parameter, ν, was estimated and MASE scores were calculated in the same manner as for the main analysis (see Sections 2.2.2 and 2.2.3 for details). Additionally, we tested fixing ν = 0.5 for all targets. All other parameters were taken from the sample specific metadata before MASE scores were calculated in the same manner as for the main analysis (see Sections 2.2.2 and 2.2.3 for details).

#### 2.2.5. Implementation

We implemented our outlier detection method and performed all evaluations in R (v4.4.1). The trend line was fitted using the `zoo` package `rollapply` function with a custom function for calculating the weighted median according to equations 2 and 3.

Loess trend lines were fitted using the `loess` function from the `stats` package. Parameters were selected to be similar to those used by estimateR [13] and ern [12]. A `span` parameter, corresponding to a window of 21 days, was used for smoothing. This method used a centrally-aligned window with decreasing weights given to the observations at the edges of the window. To fit the robust retrospective trend line the `family = “symmetric”` parameter was used with the default four iterations of a re-descending M-estimator being used with Tukey’s biweight function. While in the real-time scenario the `family = “gaussian”` parameter, for fitting by least-squares, was used instead as this is commonly done by estimateR or ern.

## 3. Results

We applied our outlier detection method to three years of digital PCR (dPCR) measurements from fourteen Swiss wastewater treatment plants (WWTPs). In the time period analysed (December 2021 - May 2025), the average frequency of outliers across viral targets ranged from 0.71 to 1.01 per 100 samples, corresponding to a yearly frequency of detected outliers of 1.55 to 2.21 outliers per year. Outlier frequency was lower for SARS and IAV than IBV and RSV. Table 1 shows outlier frequencies and assumed coefficients of pre-PCR variation (ν) for each target, which was on average 0.56 [0.46-0.64] (Figure S2).

**Table 1:**
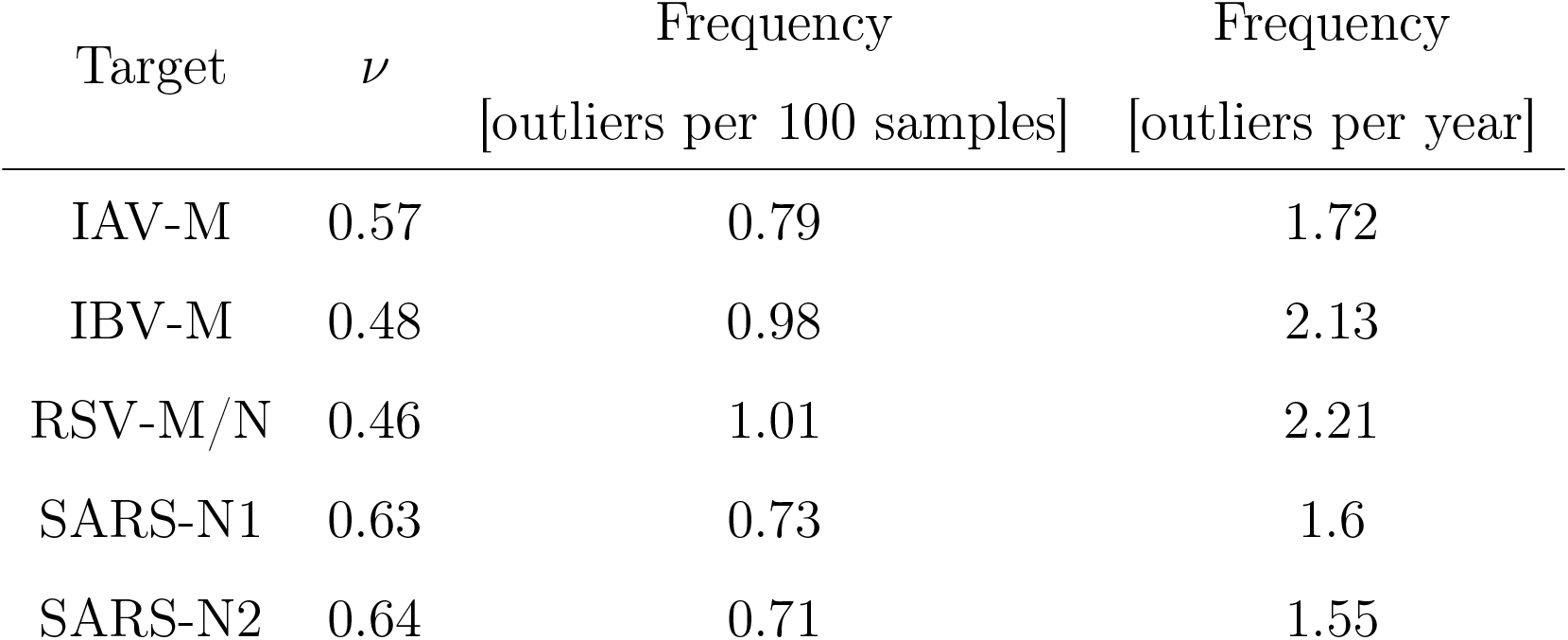
Selected pre-PCR variation (ν) parameter values for method testing and evaluation along with the corresponding frequency of outliers detected using this parameter value.

| Target | $\nu$ | Frequency<br>[outliers per 100 samples] | Frequency<br>[outliers per year] |
| --- | --- | --- | --- |
| IAV-M | 0.57 | 0.79 | 1.72 |
| IBV-M | 0.48 | 0.98 | 2.13 |
| RSV-M/N | 0.46 | 1.01 | 2.21 |
| SARS-N1 | 0.63 | 0.73 | 1.6 |
| SARS-N2 | 0.64 | 0.71 | 1.55 |

Figure 2 shows examples of the impact of outlier removal and replacement on real-time trend lines in comparison to the retrospective trend line. Generally, outlier removal and replacement brought real-time trend lines closer to the retrospective trend line (2A–D). Outliers were identified in all phases of a seasonal wave, *i.e*. during increasing (2A), decreasing (2B), or stationary (2C) concentrations in wastewater. Figure 2D highlights an important strength of our method, namely that it is able to predict outliers already with just under two weeks of historical data. However, in rare cases, the removal of outliers can bias real-time trend estimates compared to the retrospective trend. As shown in Figure 2E, our method may initially consider a dramatic increase in concentration as an outlier until further observations become available. Alternatively, if by chance a measurement is preceded by a series of low measurements, it can also be erroneously flagged as an outlier by our method (2F). We furthermore found one exception where the trend estimates were influenced by a longer gap of missing data, likely biasing not only the real-time but also the retrospective trend (Figure S3).

**Figure 2:**
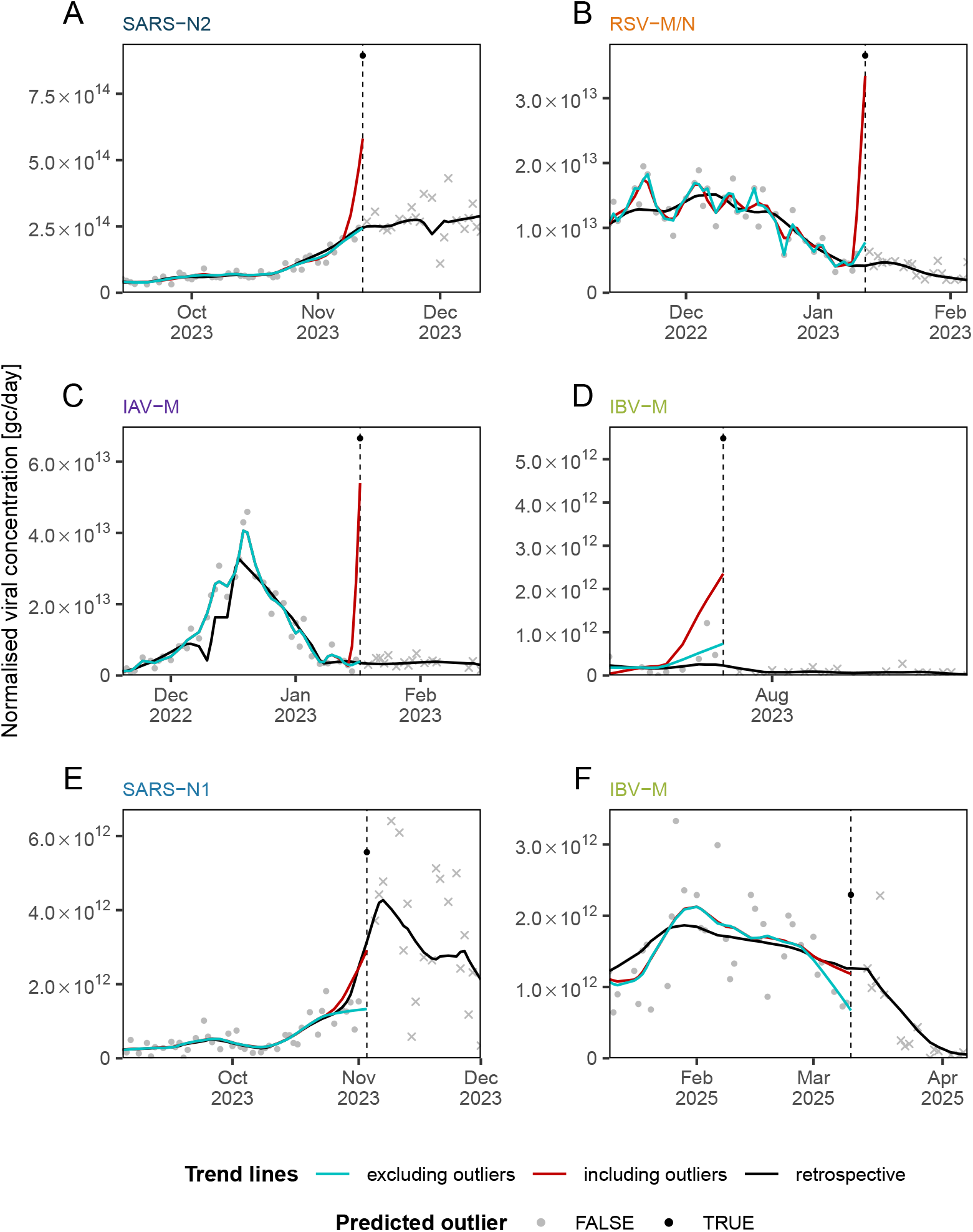
Effects of outlier removal and replacement on estimated real-time trends. Shown are real-time loess trend estimates obtained from normalised wastewater concentrations with outliers (red) and after outlier removal using our method (blue). For comparison, a retrospective trend estimate is shown in black. Observed measurements are shown as grey dots, outliers as black dots, and future measurements as grey crosses. (A-C) Real-time trend lines were improved by outlier detection when the number of infections were increasing, decreasing, or stationary. (D) Outliers were still identified when limited historical data were available. (E-F) In some cases real-time trend lines are not closer to the retrospective trend upon outlier detection, especially when there is a sharp increase in concentration that is potentially falsely identified as an outlier.

To quantify the observed effects of outlier removal and replacement on estimated real-time trends, mean absolute scaled error (MASE) scores were calculated. Their distribution over the different WWTPs for each target is shown in Figure 3. A smaller MASE score indicates closer agreement between the real-time and retrospective trend lines. When no outliers are present in a time series the MASE scores will be identical, as is the case for two time series. Outlier removal and replacement improved (*i.e*. decreased) MASE scores for all targets, most noticeably for IAV-M. Additionally, MASE scores improved for 75%, remained the same for 3%, and worsened for 22% of individual time series (Figure S4). The time series which did not show an improvement had a mean MASE score increase of 0.07, in contrast to the improved time series which had a mean decrease of 2.

**Figure 3:**
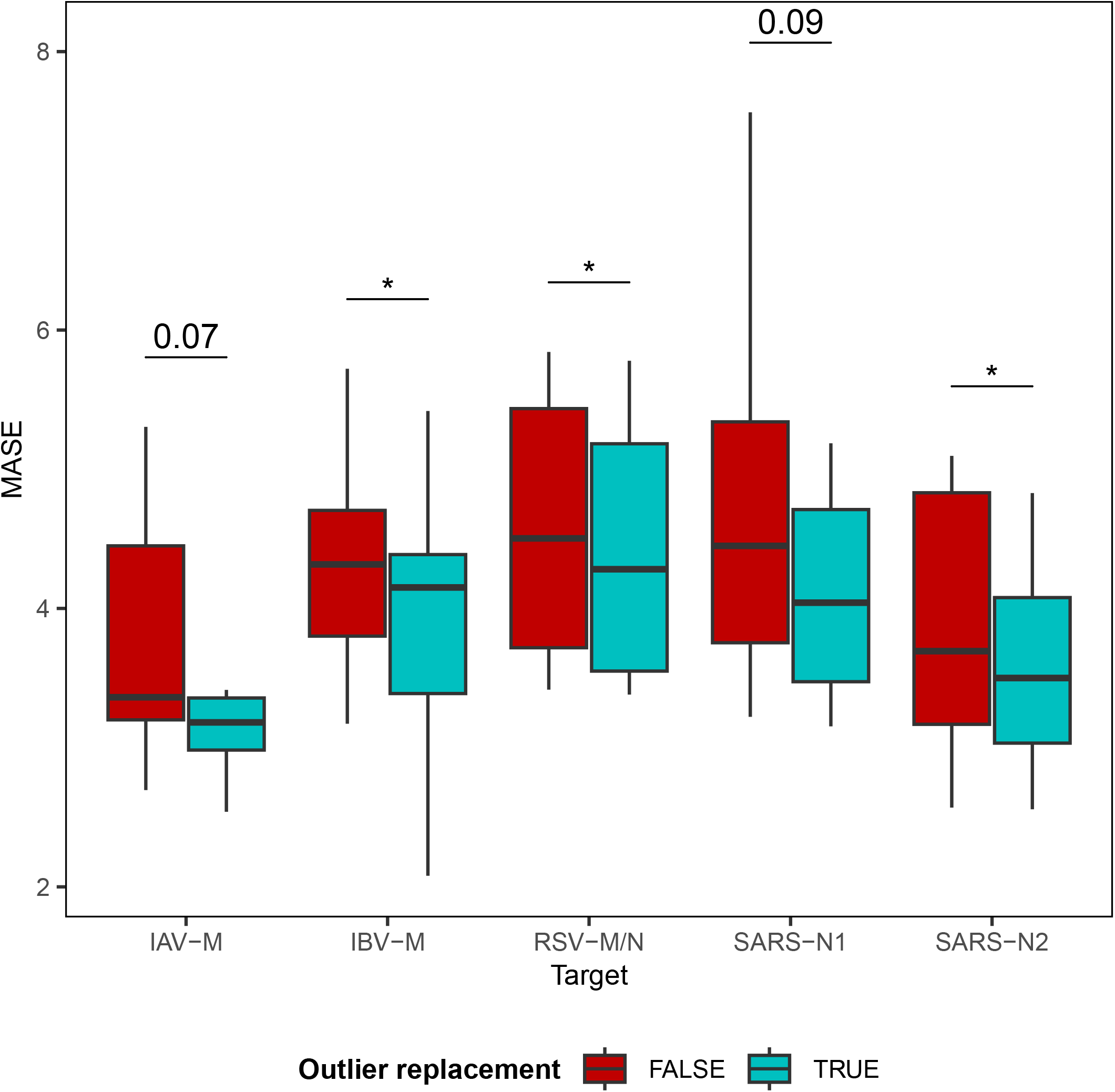
Overall time series improvement of real-time trend estimates upon outlier removal and replacement. Shown are the distributions of MASE scores over fourteen WWTPs for five different viral targets when outliers are included (red) or excluded (blue). Box plot outlier points are not shown. The sign test was used to test for consistent improvement. *: p *<* 0.05.

To assess tolerance to non-sample-specific parameters we fixed parameters to their mode or maximum value. The dilution factor, number of replicates and number of partitions were set to 30, 2 and 30,000 respectively for all samples. When doing so, the average yearly frequency of outliers ranged from 1.57 to 2.03 across viral targets and estimated ν values were all slightly higher [+0.03–0.08] compared to when per-sample specific metadata were used, most likely because less overall variation is attributed to the PCR assay when using fixed parameters (Table S2). MASE scores all decreased upon outlier removal and replacement, indicating the performance improvement remained when using these fixed parameters (Figure S5A). Additionally, MASE scores decreased upon outlier removal and replacement when ν was fixed at 0.5 for all targets (Figure S5B).

## 4. Discussion

In this paper we develop a simple method to detect outliers from a time series of pathogen concentrations in wastewater, based on digital PCR (dPCR) measurements. As in earlier work [27, 28], our method classifies outliers by their deviation from an expected trend beyond a threshold of normal variation [32]. However, we integrate an explicit model of measurement variation in dPCR to establish thresholds for outlier classification. The key strengths of our method are that it is computationally efficient, requires little historical data, and can be applied in real time.

We evaluated this approach using three years of wastewater data from fourteen wastewater treatment plants around Switzerland for four different pathogens. We found that removal and replacement of outliers detected by our method improved the robustness of real-time trend estimates, suggesting that outlier points have been correctly identified. Furthermore, this shows that real-time trend analysis is negatively influenced by extreme outliers and the improvement in real-time trend estimates upon outlier removal suggests that this is a valid approach for the handling of such data points. Real-time robustness to outliers is especially important when wastewater monitoring programmes have a mandate for timely data reporting, and reliable trend estimates can be crucial for situational awareness during an epidemic wave.

An inherent difficulty in evaluating outlier detection methods is the lack of a ground truth. We therefore evaluated our method in terms of its ability to improve the robustness of real-time trend estimates because calculated trend lines are not only displayed on monitoring dashboards but also serve as input to downstream models, e.g. for calculating wastewater-based reproduction numbers [12, 13]. Using this metric, the majority (78%) of time series were improved or unchanged. Most importantly, outlier removal always lowered the largest MASE scores to a similar range of the rest of the data (Figure S4). This highlights our method is successfully identifying the most impactful outliers. We note that differences in MASE improvements across targets likely stem from differences in the scale of outliers in the data.

Outliers were found throughout the seasonal waves, *i.e*. during stationary, increasing, and decreasing viral concentrations, suggesting their sources are likely independent from the underlying epidemic phase. The impact of correctly or incorrectly identified outliers may, however, differ across epidemic phases. For example, a false positive outlier classification at the start of a wave could delay the detection of an outbreak (Figure 2E) whereas a missed outlier early in a wave could lead to overprediction of case numbers (Figure 2A).

While we envision this method to be most useful for wastewater monitoring laboratories generating data, we wanted to consider situations where per-sample metadata may not be precisely known. We therefore investigated the impact of using fixed laboratory parameters on the outlier detection and found we were still able to effectively detect outliers. This suggests that our method can also be applied without sample-specific metadata, using fixed parameters inferred from laboratory protocols.

This method has several limitations. First, we focused our method on detecting high outlier values (*i.e*. measurements above the overall trend), as their potential impact is greater and methods to detect them rely directly on viral concentration data. This limitation likely has minimal influence on trend analysis, as low outliers are lower bounded. Additionally, low outliers are more likely caused by processes identifiable via other quality assurance methods than due to a wastewater sample randomly containing no detectable virus. This means that controlling for inhibition or extraction efficiency in the lab [23, 2], or analysing data used for signal normalisation (e.g. flow data) to monitor dilution events in the sewage system [24, 26] can help identify low outliers. Second, although our method requires less historical data than other methods, we have used historical data to estimate the level of pre-PCR variation for each target in our main analysis. Our proposed method for estimating ν would therefore only work once a sufficient amount of historical data has been collected. However, we obtained similar results when fixing pre-PCR variation to the mid point of the range across pathogens (ν = 0.5, Figure S5B), indicating our method is not highly sensitive to this parameter and can tolerate the small differences between locations and pathogens. In situations where historical data are not available, for instance when monitoring a new pathogen, similar levels of pre-PCR variation as observed for other pathogens could be assumed to estimate ν until sufficient historical data are available. Third, the standardisation of measurements in our method has been specifically tailored to dPCR data; however, our approach could be adapted to other quantification techniques, such as quantitative PCR (qPCR), which has similar characteristics. This would require estimates of the standard deviation of measurements from qPCR, which depends on qPCR-related factors such as amplification efficiency. Finally, we evaluated our method using WBE data with an average measurement frequency of four days per week. The specificity of our method is likely to decrease in less frequently sampled wastewater data. Since trends might not be so smoothly or accurately captured, there would be a greater chance of large “jumps” between observations. Outlier detection in such data sets may have to employ methods not reliant on trend line estimation. Additionally, with fewer data points the removal of outliers would be more impactful on resulting trends.

The method proposed in this work is agnostic to the underlying causes of outliers in wastewater data, which could be manifold and have not been extensively studied. In our evaluation, the removal of outliers generally improved trend estimates, suggesting that individual spikes in measured concentrations provide limited information about population-level dynamics. Possible reasons for high outliers in wastewater concentrations include individuals shedding extreme quantities of genetic material [16], non-homogeneous mixing of genetic material in the sewage [15], settling-resuspension phenomena in the sewer pipes where sedimented genetic material is washed out by extreme flow [36], and “injection” events of genetic material into the system such as dumping of a septic tank which could cause an increase in signal not related to the population in the catchment area at the time of sample collection. While our approach provides no direct insights into the origin of outliers, systematic flagging of irregular observations in wastewater data could enable more detailed analyses of such causes. For example, by sequencing wastewater samples from outlier and non-outlier days and comparing their genetic composition and diversity, host- or pathogen-related causes of outliers could be investigated. Alternatively, the association of environmental variables with outliers could be assessed, e.g. by analysing spikes in flow and rainfall data in relation to outlier and non-outlier days to investigate settling-resuspension phenomena.

Fast and reproducible outlier detection can assist researchers and public health officials when interpreting WBE data in real time. The method proposed in this work equips laboratories with a simple tool to flag potential outliers before sharing data with public health authorities and the research community, facilitating the real-time detection of potential outlier measurements for downstream analysis and visualisation on dashboards. Our results support the removal or adjusted modeling of outlier measurements, making epidemiological estimates from wastewater more reliable and improving their usefulness for public health decision-making during disease outbreaks. Together, this helps to ensure data quality in routine wastewater monitoring, which is becoming an important component of public health surveillance worldwide [37].

## Data Availability

The data used for this study are available at Zenodo: https://doi.org/10.5281/zenodo.17226976. Wastewater monitoring data generated by Eawag are publicly available at https://wisedb.ethz.ch/api/docs/.

https://doi.org/10.5281/zenodo.17226976

## Acknowledgments

We would like to thank the members of the Wastewater Monitoring Lab at Eawag for generating and making available the data used for method evaluation, with specific thanks to Melissa Pitton, Charlie Gan, Daniela Yordanova, Jolinda de Korne and Patrick Schmidhalter for their input. Additionally, we thank members of the WISE (Wastewater-based Infectious disease Surveillance and Epidemiology) project for their support and intellectual contributions.

## Authors’ contributions

REM: Conceptualisation, Methodology, Formal Analysis, Writing - Original Draft, Visualisation. TRJ: Writing - Review & Editing, Supervision, Funding acquisition. TS: Writing - Review & Editing, Supervision, Funding acquisition. AL: Conceptualisation, Methodology, Writing - Original Draft, Supervision.

## Appendix A. Dilution factor calculation

For our protocol the dilution factor was calculated as

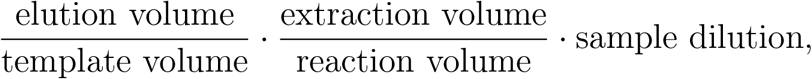

where extraction volume is the volume of wastewater taken for extraction (40,000 µL); elution volume is the total volume of RNA extract after processing (80 µL); template volume is the volume of RNA extract added to the dPCR reaction (5 µL); reaction volume is the total volume of RNA template and reagents in the dPCR reaction (25 µL); and sample dilution is a factor of how much the RNA extract was diluted with water to reduce inhibition (WWTP specific, 3 or 5).

## Appendix B. Supplementary Materials

**Table S1:** Details of the wastewater treatment plants in the monitoring programme. 10 treatment plants are still being monitored.

| Wastewater<br>Treatment Plant | Main<br>Town | Catchment<br>Population | Sampling<br>Start | Sampling<br>End |
| --- | --- | --- | --- | --- |
| ARA Altenrhein | Altenrhein | 64000 | 01/12/2021 | NA |
| ARA Basel/Prorheno | Basel | 268317 | 10/07/2023 | NA |
| ARA Buholz | Luzern | 178555 | 10/07/2023 | NA |
| ARA Chur | Chur | 55000 | 01/12/2021 | NA |
| ARA Region Bern | Bern | 224557 | 10/07/2023 | 01/01/2025 |
| ARA Schwyz | Schwyz | 31164 | 10/07/2023 | 01/01/2025 |
| ARA Sensetal | Laupen | 62000 | 01/12/2021 | NA |
| ARA Werdhoelzli | Zurich | 471000 | 01/12/2021 | NA |
| ARA Zuchwil | Solothurn | 95793 | 10/07/2023 | 01/01/2025 |
| CDA Lugano | Lugano | 124000 | 01/12/2021 | NA |
| STEP Aire | Geneva | 454000 | 01/12/2021 | NA |
| STEP Neuchatel | Neuchatel | 40697 | 10/07/2023 | NA |
| STEP Porrentruy | Porrentruy | 16500 | 10/07/2023 | 01/01/2025 |
| STEP Vidy | Lausanne | 240000 | 10/07/2023 | NA |

**Table S2:** Selected pre-PCR variation (ν) parameter values for sensitivity analysis along with the corresponding frequency of outliers detected with this value.

| Target | $\nu$ | Frequency | Frequency |
| --- | --- | --- | --- |
|  |  | [outliers per 100 samples] | [outliers per year] |
| IAV-M | 0.62 | 0.78 | 1.69 |
| IBV-M | 0.55 | 0.93 | 2.03 |
| RSV-M/N | 0.54 | 0.93 | 2.03 |
| SARS-N1 | 0.66 | 0.74 | 1.61 |
| SARS-N2 | 0.67 | 0.72 | 1.57 |

**Figure S1:**
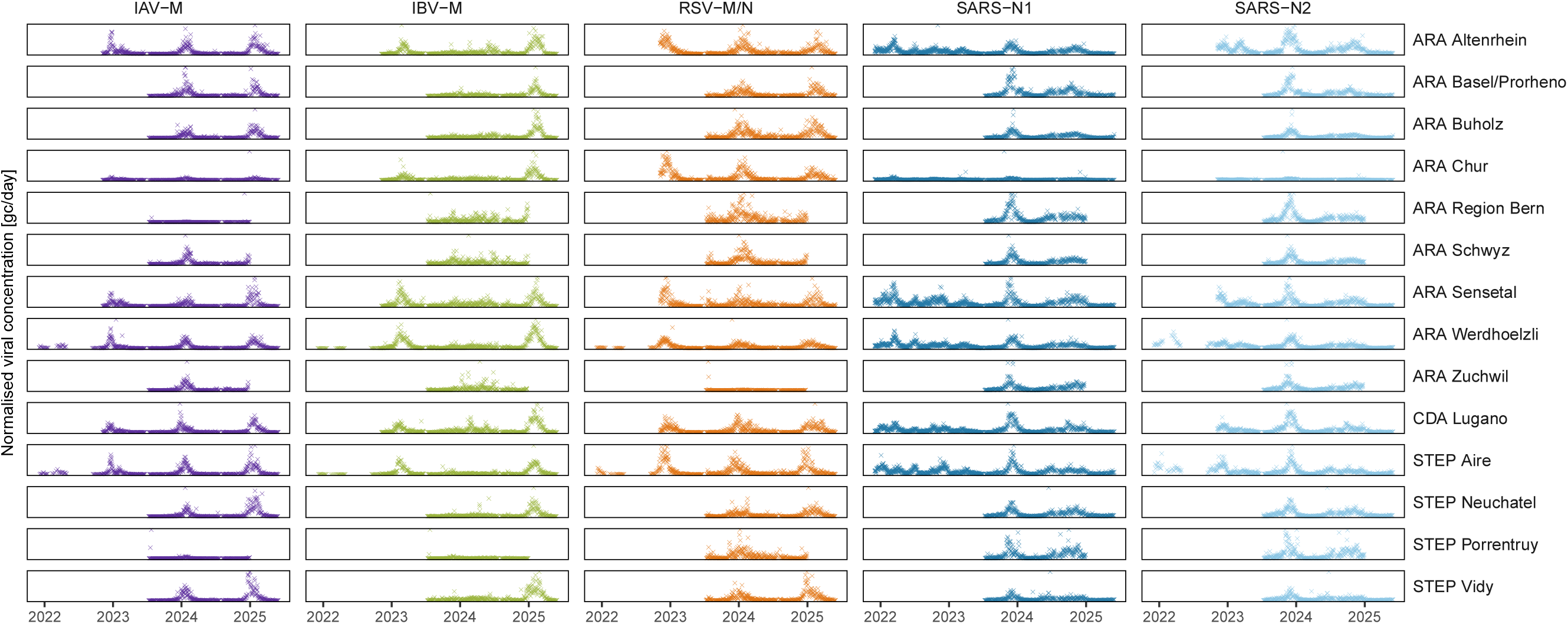
Overview of the different time series

**Figure S2:**
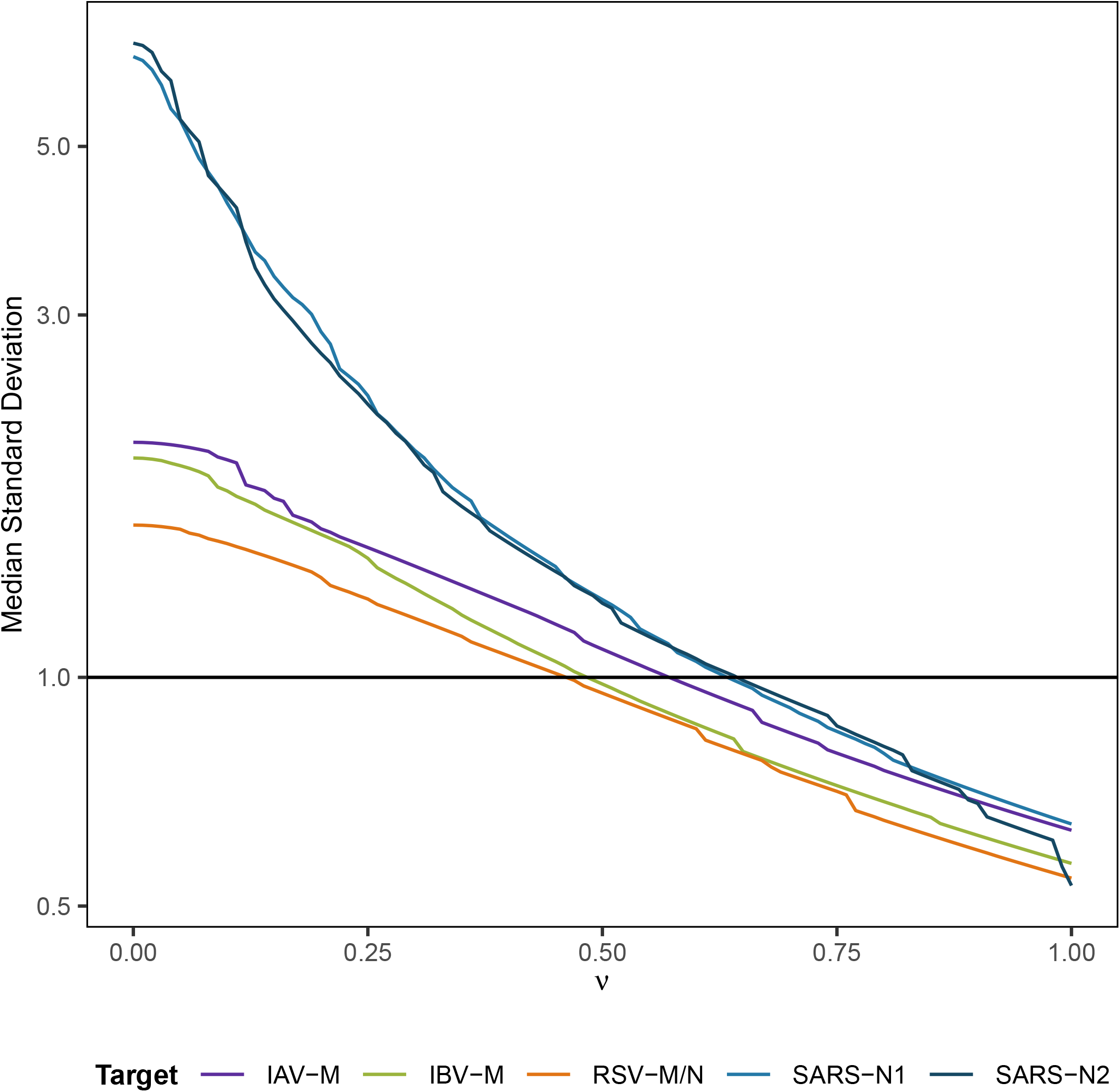
Median standard deviation over the different wastewater treatment plants for different values of ν. It is plotted separately for each of the viral targets (different colours).

**Figure S3:**
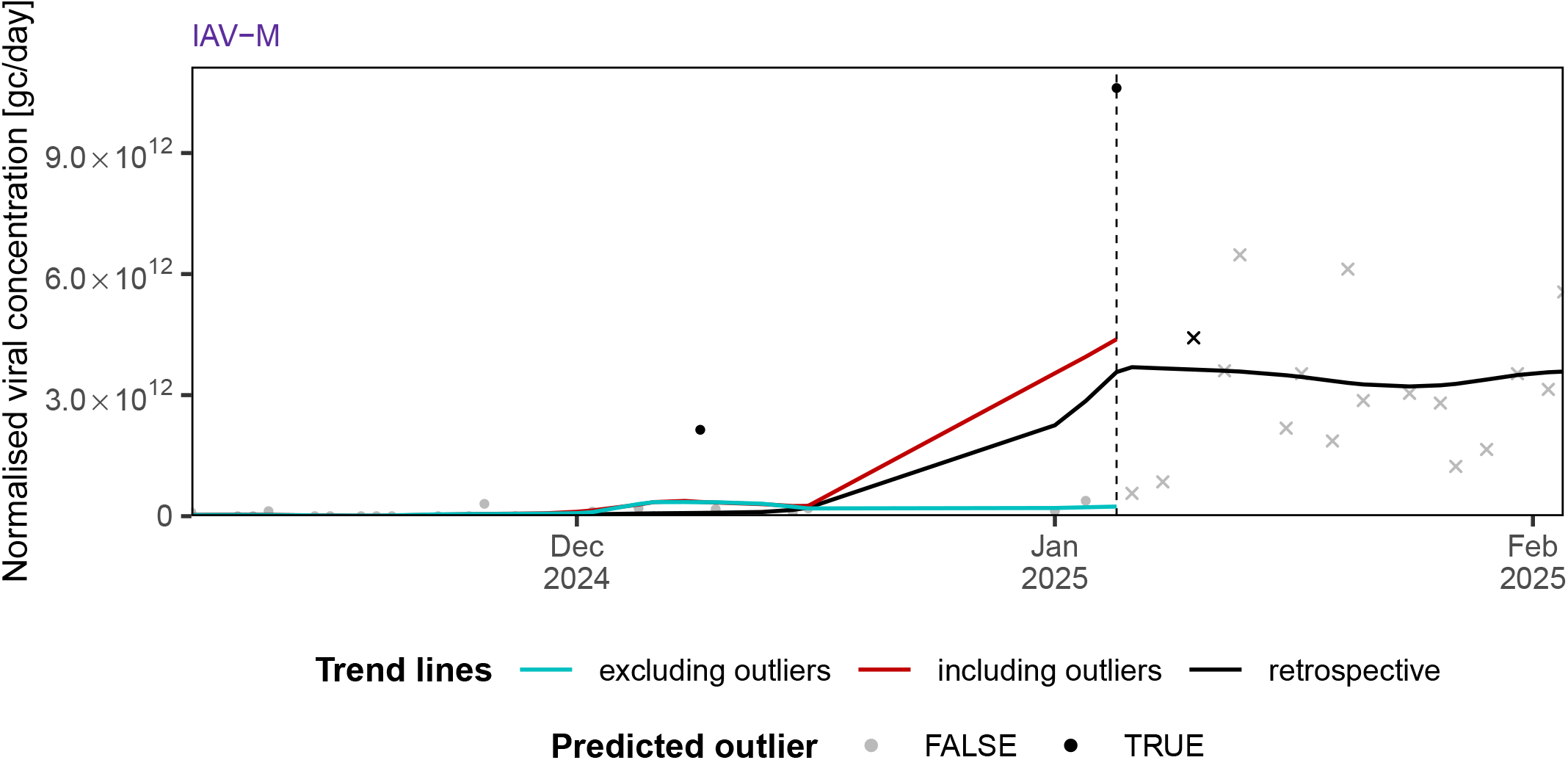
In one observed case the real-time without removal of outliers (red) was closer to the retrospective trend (black) than real-time with removal (blue). Observed measurements are shown as grey dots, outliers as black dots, and future measurements as grey crosses. The gap of missing data (mid-Dec - Jan) has likely impacted the robustness of the retrospective trend.

**Figure S4:**
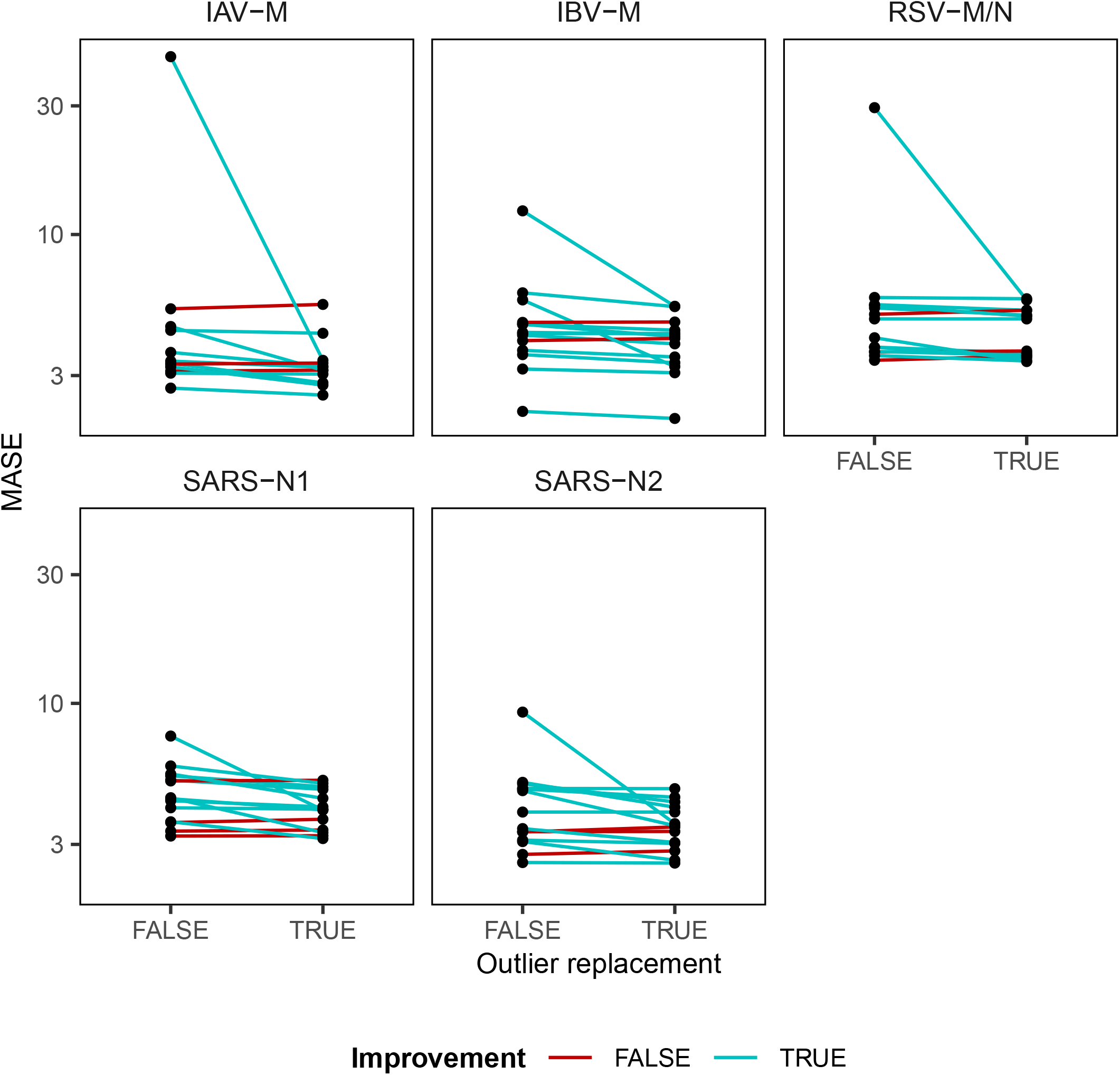
Paired differences in MASE scores with and without outliers. Shown are semi-log plots of MASE scores, paired for each WWTP and separated by target (different panels). When outlier removal decreases MASE scores we have an improvement (blue) in contrast to an increase in MASE scores (red).

**Figure S5:**
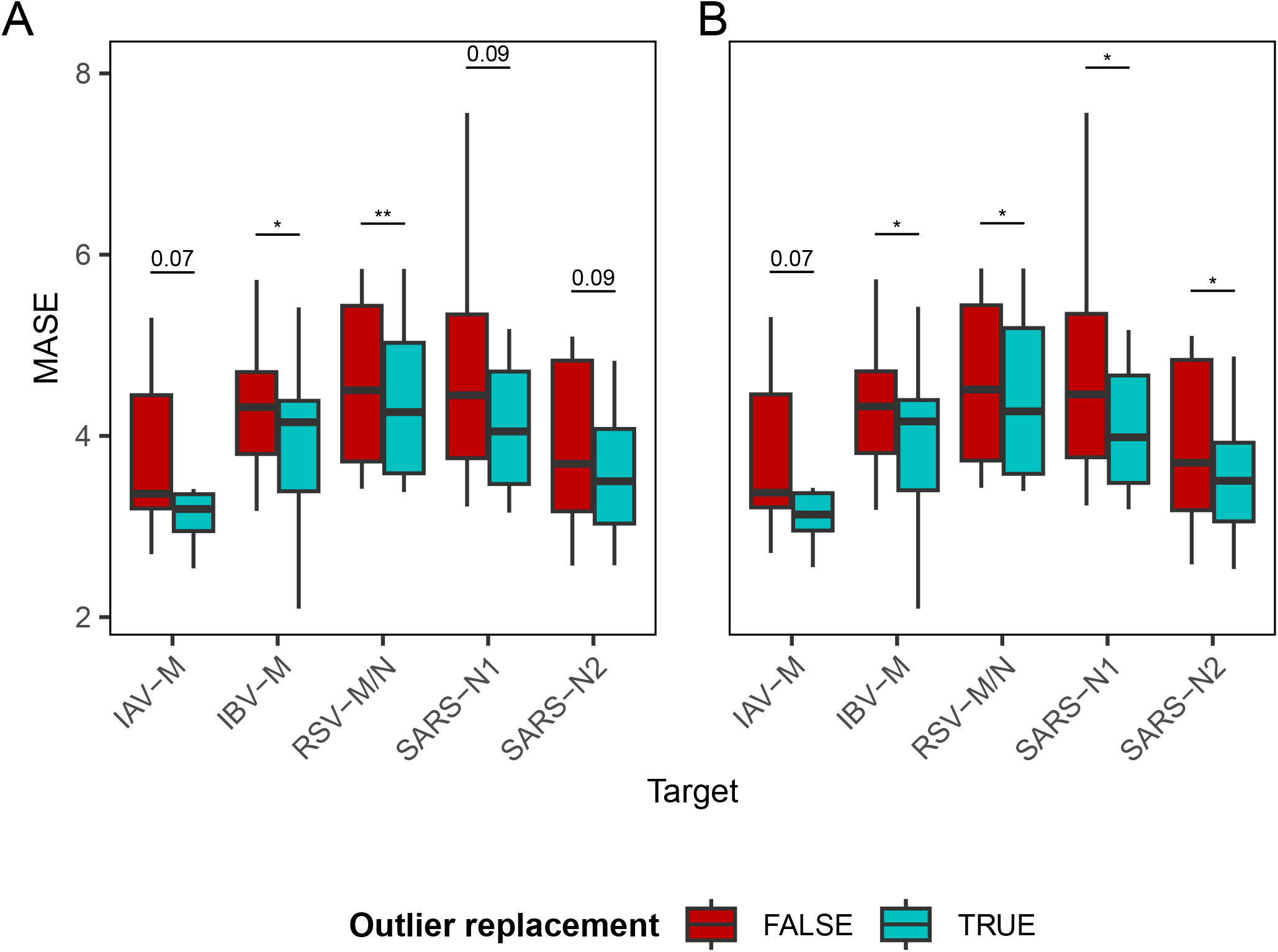
Overall time series improvement of real-time trend estimates upon outlier removal and replacement when either (A) lab parameters were fixed at their most common/maximal value or (B) pre-PCR variation was fixed at 0.5. Shown are the distributions of MASE scores over fourteen WWTPs for five different viral targets when outliers are included (red) or excluded (blue). Box plot outlier points are not shown. The sign test was used to test for consistent improvement. *: p < 0.05, **; p < 0.01

## References

[1] D. A. Bowes, Towards a Precision Model for Environmental Public Health: Wastewater-based Epidemiology to Assess Population-level Exposures and Related Diseases, Current Epidemiology Reports (Jun. 2024). doi:10.1007/s40471-024-00350-6.

[2] M. Pitton, R. E. McLeod, L. Caduff, A. Dauletova, J. de Korne-Elenbaas, C. Gan, C. Hablützel, A. Holschneider, S. Kang, G. Loustalot, P. Schmidhalter, L. Schneider, A. Wettlauffer, D. Yordanova, T. R. Julian, C. Ort, A six-plex digital PCR assay for monitoring respiratory viruses in wastewater (Dec. 2024). doi:10.1101/2024.12.06.24317241.

[3] A. B. Boehm, B. Hughes, D. Duong, V. Chan-Herur, A. Buchman, M. K. Wolfe, A. J. White, Wastewater concentrations of human influenza, metapneumovirus, parainfluenza, respiratory syncytial virus, rhinovirus, and seasonal coronavirus nucleic-acids during the COVID-19 pandemic: A surveillance study, The Lancet Microbe 4 (5) (2023) e340–e348. doi:10.1016/S2666-5247(22)00386-X.

[4] G. Medema, L. Heijnen, G. Elsinga, R. Italiaander, A. Brouwer, Presence of SARS-Coronavirus-2 RNA in Sewage and Correlation with Reported COVID-19 Prevalence in the Early Stage of the Epidemic in The Netherlands, Environmental Science & Technology Letters 7 (7) (2020) 511–516. doi:10.1021/acs.estlett.0c00357.

[5] A. Rabe, S. Ravuri, E. Burnor, J. A. Steele, R. S. Kantor, S. Choi, S. Forman, R. Batjiaka, S. Jain, T. M. León, D. J. Vugia, A. T. Yu, Correlation between wastewater and COVID-19 case incidence rates in major California sewersheds across three variant periods, Journal of Water and Health 21 (9) (2023) 1303–1317. doi:10.2166/wh.2023.173.

[6] J. S. Huisman, J. Scire, L. Caduff, X. Fernandez-Cassi, P. Ganesanandamoorthy, A. Kull, A. Scheidegger, E. Stachler, A. B. Boehm, B. Hughes, A. Knudson Topol, K. R. Wigginton, M. K. Wolfe, T. Kohn, C. Ort, T. Stadler, T. R. Julian, Wastewater-Based Estimation of the Effective Reproductive Number of SARS-CoV-2, Environmental Health Perspectives 130 (5) (2022) 057011. doi: 10.1289/EHP10050.

[7] Bagutti, M. A. Hug, P. Heim, L. M. Pekerman, E. I. Hampe, P. Hübner, S. Fuchs, M. Savic, T. Stadler, I. Topolsky, P. I. Baykal, D. Dreifuss, N. Beerenwinkel, S. T. Sutter, Wastewater monitoring of SARS-CoV-2 shows high correlation with COVID-19 case numbers and allowed early detection of the first confirmed B.1.1.529 infection in Switzerland: Results of an observational surveillance study, Swiss Medical Weekly 152 (2526) (2022) w30202–w30202. doi:10.4414/SMW.2022.w30202.

[8] H. Schenk, P. Heidinger, H. Insam, N. Kreuzinger, R. Markt, F. Nägele, H. Oberacher, C. Scheffknecht, M. Steinlechner, G. Vogl, A. O. Wagner, W. Rauch, Prediction of hospitalisations based on wastewater-based SARS-CoV-2 epidemiology, Science of The Total Environment 873 (2023) 162149. doi:10.1016/j.scitotenv.2023.162149.

[9] A. Galani, R. Aalizadeh, M. Kostakis, A. Markou, N. Alygizakis, T. Lytras, P. G. Adamopoulos, J. Peccia, D. C. Thompson, A. Kontou, A. Karagiannidis, E. S. Lianidou, M. Avgeris, D. Paraskevis, S. Tsiodras, A. Scorilas, V. Vasiliou, M.-A. Dimopoulos, N. S. Thomaidis, SARS-CoV-2 wastewater surveillance data can predict hospitalizations and ICU admissions, Science of The Total Environment 804 (2022) 150151. doi:10.1016/j.scitotenv.2021.150151.

[10] Y. Okada, H. Nishiura, Estimating the effective reproduction number of COVID-19 from population-wide wastewater data: An application in Kagawa, Japan, Infectious Disease Modelling 9 (3) (2024) 645–656. doi:10.1016/j.idm.2024.03.006.

[11] S. Nadeau, A. J. Devaux, C. Bagutti, M. Alt, E. I. Hampe, M. Kraus, E. Würfel, K. N. Koch, S. Fuchs, S. Tschudin-Sutter, A. Holschneider, C. Ort, C. Chen, J. S. Huisman, T. R. Julian, T. Stadler, Influenza transmission dynamics quantified from RNA in wastewater in Switzerland, Swiss Medical Weekly 154 (1) (2024) 3503–3503. doi:10.57187/s.3503.

[12] Champredon, I. Papst, W. Yusuf, Ern: An R package to estimate the effective reproduction number using clinical and wastewater surveillance data, PLOS ONE 19 (6) (2024) e0305550. doi:10.1371/journal.pone.0305550.

[13] J. Scire, J. S. Huisman, A. Grosu, D. C. Angst, A. Lison, J. Li, M. H. Maathuis, S. Bonhoeffer, T. Stadler, estimateR: An R package to estimate and monitor the effective reproductive number, BMC Bioinformatics 24 (1) (2023) 310. doi: 10.1186/s12859-023-05428-4.

[14] A. Lison, Adrian-lison/EpiSewer: EpiSewer 0.0.1, Zenodo (Jan. 2024). doi: 10.5281/zenodo.10569102.

[15] M. J. Wade, A. Lo Jacomo, E. Armenise, M. R. Brown, J. T. Bunce, G. J. Cameron, Z. Fang, K. Farkas, D. F. Gilpin, D. W. Graham, J. M. S. Grimsley, A. Hart, T. Hoffmann, K. J. Jackson, D. L. Jones, C. J. Lilley, J. W. McGrath, J. M. McKinley, C. McSparron, B. F. Nejad, M. Morvan, M. Quintela-Baluja, A. M. I. Roberts, A. C. Singer, C. Souque, V. L. Speight, C. Sweetapple, D. Walker, G. Watts, A. Weightman, B. Kasprzyk-Hordern, Understanding and managing uncertainty and variability for wastewater monitoring beyond the pandemic: Lessons learned from the United Kingdom national COVID-19 surveillance programmes, Journal of Hazardous Materials 424 (2022) 127456. doi:10.1016/j.jhazmat.2021.127456.

[16] X. Li, S. Zhang, J. Shi, S. P. Luby, G. Jiang, Uncertainties in estimating SARS-CoV-2 prevalence by wastewater-based epidemiology, Chemical Engineering Journal 415 (2021) 129039. doi:10.1016/j.cej.2021.129039.

[17] R. Arabzadeh, D. M. Grünbacher, H. Insam, N. Kreuzinger, R. Markt, W. Rauch, Data filtering methods for SARS-CoV-2 wastewater surveillance, Water Science and Technology 84 (6) (2021) 1324–1339. doi:10.2166/wst.2021.343.

[18] The European Wastewater Surveillance Dashboard, https://arcgis.jrc.ec.europa.eu/portal/apps/dashboards/e296cdf0c0d042e6b60b07a351f2dc5c (Mar. 2025).

[19] ESR Wastewater Surveillance, https://www.poops.nz/ (Mar. 2025).

[20] WastewaterSCAN Dashboard, https://data.wastewaterscan.org/ (Mar. 2025).

[21] Z. Fang, A. M. I. Roberts, C.-D. Mayer, A. Frantsuzova, J. M. Potts, G. J. Cameron, P. T. R. Singleton, I. Currie, Wastewater monitoring of COVID-19: A perspective from Scotland, Journal of Water and Health 20 (12) (2022) 1688–1700. doi:10.2166/wh.2022.082.

[22] H. Schenk, W. Rauch, A. Zulli, A. B. Boehm, SARS-CoV-2 surveillance in US wastewater: Leading indicators and data variability analysis in 2023–2024, PLOS ONE 19 (11) (2024) e0313927. doi:10.1371/journal.pone.0313927.

[23] The dMIQE Group, J. F. Huggett, The Digital MIQE Guidelines Update: Minimum Information for Publication of Quantitative Digital PCR Experiments for 2020, Clinical Chemistry 66 (8) (2020) 1012–1029. doi:10.1093/clinchem/hvaa125.

[24] W. Rauch, H. Schenk, H. Insam, R. Markt, N. Kreuzinger, Data modelling recipes for SARS-CoV-2 wastewater-based epidemiology, Environmental Research 214 (2022) 113809. doi:10.1016/j.envres.2022.113809.

[25] R. Markt, L. Endler, F. Amman, A. Schedl, T. Penz, M. Büchel-Marxer, Grünbacher, M. Mayr, E. Peer, M. Pedrazzini, W. Rauch, A. O. Wagner, Allerberger, A. Bergthaler, H. Insam, Detection and abundance of SARS-CoV-2 in wastewater in Liechtenstein, and the estimation of prevalence and impact of the B.1.1.7 variant, Journal of Water and Health 20 (1) (2021) 114–125. doi:10.2166/wh.2021.180.

[26] A. Lastra, J. Botello, A. Pinilla, J. I. Urrutia, J. Canora, J. Sánchez, P. Fernández, F. J. Candel, A. Zapatero, M. Ortega, J. Flores, SARS-CoV-2 detection in wastewater as an early warning indicator for COVID-19 pandemic. Madrid region case study, Environmental Research 203 (2022) 111852. doi: 10.1016/j.envres.2021.111852.

[27] Klaassen, R. H. Holm, T. Smith, T. Cohen, A. Bhatnagar, N. A. Menzies, Predictive power of wastewater for nowcasting infectious disease transmission: A retrospective case study of five sewershed areas in Louisville, Kentucky, Environmental Research 240 (2024) 117395. doi:10.1016/j.envres.2023.117395.

[28] D. G. Manuel, G. Saran, I. Lee, W. Yusuf, M. Thomson, É. Mercier, V. Pileggi, R. M. McKay, R. Corchis-Scott, Q. Geng, M. Servos, H. Inert, H. Dhiyebi, I. M. Yang, B. Harvey, E. Rodenburg, C. Millar, R. Delatolla, Wastewater-based surveillance of SARS-CoV-2: Short-term projection (forecasting), smoothing and outlier identification using Bayesian smoothing, Science of The Total Environment 949 (2024) 174937. doi:10.1016/j.scitotenv.2024.174937.

[29] M. Courbariaux, N. Cluzel, S. Wang, V. Maréchal, L. Moulin, S. Wurtzer,O. Consortium, J.-M. Mouchel, Y. Maday, G. Nuel, I. Bertrand, M. Boni, Gantzer, S. F. Le Guyader, Y. Maday, V. Maréchal, J.-M. Mouchel, L. Moulin, R. Teyssou, S. Wurtzer, A Flexible Smoother Adapted to Censored Data With Outliers and Its Application to SARS-CoV-2 Monitoring in Wastewater, Frontiers in Applied Mathematics and Statistics 8 (Feb. 2022). doi:10.3389/fams.2022.836349.A

[30] A. L. Rainey, S. Liang, J. H. B. Jr, T. Sabo-Attwood, A. T. Maurelli, A multistate assessment of population normalization factors for wastewater-based epidemiology of COVID-19, PLOS ONE 18 (4) (2023) e0284370. doi:10.1371/journal.pone.0284370.

[31] Lison, T. R. Julian, T. Stadler, Improving inference in wastewater-based epidemiology by modelling the statistical features of digital PCR (Oct. 2024). doi:10.1101/2024.10.14.618307.

[32] V. Hodge, J. Austin, A Survey of Outlier Detection Methodologies, Artificial Intelligence Review 22 (2) (2004) 85–126. doi:10.1023/B:AIRE.0000045502.10941.a9.

[33] A. S. Yaro, F. Maly, P. Prazak, K. Malý, Outlier Detection Performance of a Modified Z-Score Method in Time-Series RSS Observation With Hybrid Scale Estimators, IEEE Access 12 (2024) 12785–12796. doi:10.1109/ACCESS.2024.3356731.

[34] WISE, https://wise.ethz.ch/ (Aug. 2024).

[35] W. S. Cleveland, E. Grosse, W. M. Shyu, Local Regression Models, in: Statistical Models in S, Routledge, 1992, p. Chapter 8.

[36] A.-M. Solvi, Modelling the sewer-treatment-urban river system in view of the EU Water Framework Directive, PhD (2007).

[37] C. Naughton, F. A. Roman, Jr, A. G. F. Alvarado, A. Q. Tariqi, M. A. Deeming, K. F. Kadonsky, K. Bibby, A. Bivins, G. Medema, W. Ahmed, P. Katsivelis, V. Allan, R. Sinclair, J. B. Rose, Show us the data: Global COVID-19 wastewater monitoring efforts, equity, and gaps, FEMS Microbes 4 (2023) xtad003. doi:10.1093/femsmc/xtad003.

